# Comparative Effectiveness of the Bivalent (Original/Omicron BA.4/BA.5) mRNA COVID-19 Vaccines mRNA-1273.222 and BNT162b2 Bivalent in Adults in the United States

**DOI:** 10.1101/2023.07.12.23292576

**Authors:** Hagit Kopel, Van Hung Nguyen, Catherine Boileau, Alina Bogdanov, Isabelle Winer, Thierry Ducruet, Ni Zeng, Mac Bonafede, Daina B. Esposito, David Martin, Andrew Rosen, Nicolas Van de Velde, Sten H Vermund, Stefan Gravenstein, James A. Mansi

**Author notes:** Corresponding author: James A. Mansi. Joint first authors (Hagit Kopel, Van Hung Nguyen).

## Abstract

**Background:** The emergence of Omicron variants coincided with declining vaccine-induced protection against SARS-CoV-2 infection and other COVID-19-related outcomes. Two bivalent mRNA vaccines, mRNA-1273.222 (Moderna) and BNT162b2 Bivalent (Pfizer-BioNTech) were developed to provide greater protection against the predominate circulating variants by including the mRNA that encodes both the ancestral (original) strain and BA.4/BA.5. We estimated their relative vaccine effectiveness (rVE) in preventing COVID-19-related outcomes in the US.

**Methods:** We conducted a retrospective cohort study using a US nationwide dataset linking primary care electronic health records (EHR) and pharmacy/medical claims data. The adult study population (aged ≥18 years) received either mRNA-1273.222 or BNT162b2 Bivalent vaccination between August 31, 2022, and February 28, 2023. We used a propensity score weighting based on the inverse probability of treatment to adjust for the baseline differences in age, sex, race, ethnicity, geographic region, vaccination week, and health status between groups. Outcomes evaluated were rVE of the two bivalent mRNA vaccines against COVID-19-related hospitalizations (primary outcome) and outpatient visits (secondary). We weighted the vaccine groups prior to analysis and estimated adjusted hazard ratios (HR) using multivariable Cox regression models. We calculated rVE as (1−HR) × 100.

**Results:** We evaluated outcomes for 1,034,538 mRNA-1273.222 and 1,670,666 BNT162b2 Bivalent vaccine recipients. The adjusted rVE of mRNA-1273.222 versus BNT162b2 Bivalent vaccines against COVID-19-related hospitalization was 9.8% (95% confidence interval: 2.6%–16.4%). The adjusted rVE against COVID-19-related outpatient visits was 5.1% (95% CI: 3.2%–6.9%). When evaluated by age group, the incremental relative effectiveness was greater. Among adults ≥ 65, rVE against COVID-19-related hospitalizations and outpatient visits was 13.5% (95% CI: 5.5%–20.8%) and 10.7% (8.2%–13.1%), respectively.

**Conclusion:** We found greater effectiveness of mRNA-1273.222 compared with the BNT162b2 Bivalent vaccine in preventing COVID-19-related hospitalizations and outpatient visits, with increased benefits in older adults.

## Introduction

Despite high levels of vaccine effectiveness (VE) against the ancestral SARS-CoV-2 strain and variants observed in the early stages of the COVID-19 pandemic, the VE of a primary series of mRNA-1273 or BNT162b2 monovalent vaccines decreased with the emergence of the Delta and Omicron (B.1.1.529 [BA.1]) variants and Omicron’s subvariants (e.g. BA.2, BA.4, BA.5, XBB.1)^1, 2^. Administration of a monovalent vaccine booster dose improved both immunogenicity and VE; however, effectiveness remained lower compared to that observed against earlier circulating SARS-CoV-2 variants. This led to March 2022 recommendations of yet an additional booster dose for vulnerable individuals, highlighting concern for waning protection and aiming to reduce disease severity and frequency of breakthrough infection.^3–7^.

To counter waning immunity and broaden protection against emerging variants, new bivalent mRNA vaccines were developed targeting both the spike protein of the Omicron BA.4/BA.5 subvariant and the ancestral (original) SARS-CoV-2. These bivalent vaccines demonstrated increased protection against COVID-19-related symptomatic infections and severe outcomes.^1, 8–12^ and substantially boosted clinical protection and antibody titers for up to 6 months.^13^ While the safety profile was similar between monovalent and bivalent mRNA vaccines, greater immunogenicity and relative vaccine effectiveness (rVE) were demonstrated with increased time since prior infection or prior COVID-19 vaccination.^12, 14–22^ On August 31, 2022,^23^ bivalent vaccines (Original/ Omicron BA.4/BA.5) were authorized in the United States (US) by the Food and Drug Administration (FDA) as a booster dose for adults (≥18) and are now recommended by the US Centers for Disease Control and Prevention (CDC) for all individuals ≥6 months of age.^24^ However, while approximately 70% of the eligible US population has completed a primary series, only ∼17% of those eligible have received bivalent vaccination as of June 2023.^25^

While hospitalization rates during the period of omicron predominance are lower than those observed earlier in the pandemic,^25^ more vulnerable sub-groups, including unvaccinated adults, older adults, immunocompromised persons, and those with certain chronic underlying medical conditions are at increased risk for severe disease outcomes and death. Hence, vaccine effectiveness and waning protection over time has immediate public health implications.^26^ Furthermore, this information is essential for COVID-19 vaccine decision-makers and health care providers (HCP), to provide and convey presumptive recommendations and increase vaccine confidence among patients, particularly those at higher risk for COVID-19-related morbidity and mortality. ^25, 27, 28^

Previous analyses of the primary series and booster reported slightly higher vaccine effectiveness for the monovalent mRNA-1273 (Moderna) vaccine compared with BNT162b2 (Pfizer-BioNTech) in preventing both reported cases and COVID-19-related hospitalizations and outpatients visits. ^29–32^ However, rVE of these bivalent mRNA vaccines has not been established. The current study evaluated data from a large nationwide dataset to estimate the rVE of the mRNA-1273.222 bivalent vaccine (Original/ Omicron BA.4/BA.5) with BNT162b2 Bivalent vaccine (Original/ Omicron BA.4/BA.5) administered for the prevention of COVID-19-related hospitalizations and outpatient visits in adults in the US.

## Methods

### Data source and de-identification

We performed an observational, retrospective cohort study using real world data from two integrated sources. Widely used primary care electronic health record (EHR) platforms in the US (the Veradigm EHR dataset includes the Allscripts Tier 1, Allscripts Tier 2, and Practice Fusion EHR) served as the first source. Pharmacy and medical claims data (Komodo dataset) served as the second source. Past research successfully used this integrated dataset to evaluate vaccine effectiveness^32–40^ and its data to contain key variables for conducting effectiveness /outcomes research and demonstrated it to be representative of the US population. The dataset detailed by Nguyen et al 2023 includes variables based on structured data (e.g. from ICD-10 codes) across all care settings^32^. Each data asset meets the minimum protected health information (PHI) data requirements as specified for use by the algorithm. Research staff were not involved in preparation of datasets containing PHI or the actual running of the algorithm. The final linked data set merged the patient-level de-identified tokens in each individual dataset and contained no PHI. This linked, de-identified dataset was compliant with the Health Insurance Portability and Accountability Act (HIPAA) and certified for research use. The retrospective observational study was designed, implemented, and reported in accordance with Good Pharmacoepidemiological Practice, applicable local regulations, and the ethical principles laid down in the Declaration of Helsinki. Approval by an institutional review board and patient consent were not necessary as the study was a noninterventional, retrospective cohort using a certified HIPAA-compliant deidentified research database.

In this study, we have used closed claims in the main analysis and open claims in the sensitivity analysis. The closed claims dataset follows three-tiered specified requirements (see **Supplementary Materials**). Open dataset includes all available claims for a patient, regardless of enrollment period, payer source, or closure status.

### Participants and study design

Adults ≥18 years of age who had received either the mRNA-1273.222 (50 mcg), or BNT162b2 Bivalent vaccine (30 mcg) between August 31, 2022, and February 28, 2023, were eligible for inclusion in the study (see **Supplementary Table 1**). We defined the index date as the vaccination date with the bivalent vaccine, and the cohort entry date (CED) as 7 days later (**Figure 1**). Through the databases, we followed-up individuals for outcomes of interest from the CED until the end of the available data (February 28, 2023), receipt of another COVID-19 vaccine, or disenrollment from their medical/pharmacy plan, whichever occurred first. We classified individuals into two exposure cohorts based on the type of bivalent vaccine received (see **Supplementary Table 1** for a list of codes used to identify vaccinated individuals), with each individual only included once.

**Figure 1.**
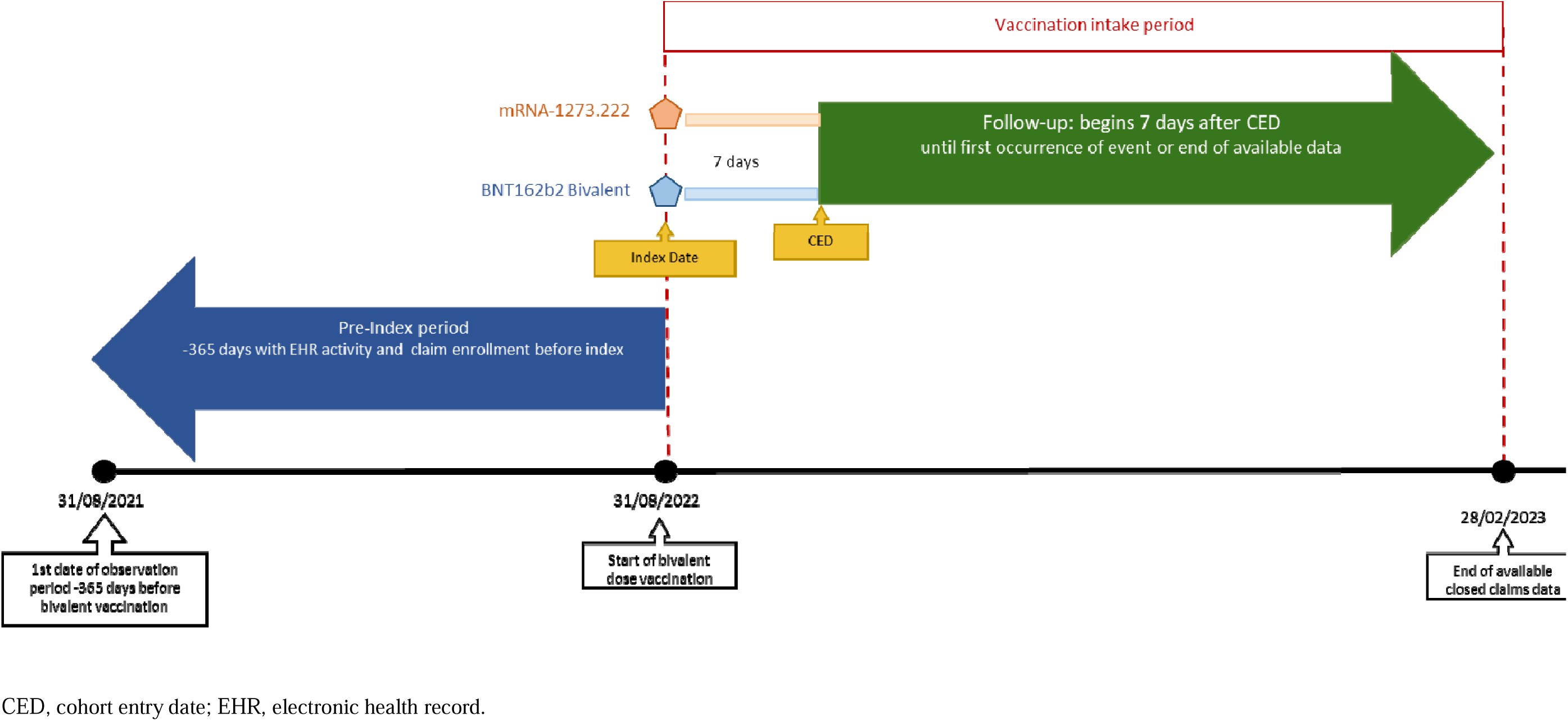
Study design.

Inclusion criteria included a minimum of 365 days of continuous medical and pharmacy enrollment in a health plan contributing data to the claim set prior to receipt of the booster (index date), and at least one contact with a health service provider in the 365 days prior to the index date. Exclusion criteria included evidence of SARS-CoV-2 infection or vaccination within the first 7 days after receipt of the bivalent vaccine, less than 1 day of follow-up, or missing birth year or sex.

### Study objectives

The primary objective of this study was assessment of the rVE between mRNA-1273.222 and BNT162b2 Bivalent in preventing COVID-19-related illness requiring hospitalization, evaluated from the cohort entry date to first occurrence. The secondary objective was assessment of the rVE in prevention of COVID-19-related outpatient visits, evaluated from cohort entry date to first occurrence. We identified inpatient admissions based on all claims billed in the inpatient setting. Among these claims, we assigned those associated with inpatient hospital services to the inpatient hospital designation, and those associated with non-inpatient services (such as skilled nursing facilities or emergency department visits) to the outpatient designation. Once encounters were identified as either inpatient or outpatient, we flagged claims with a diagnosis code for COVID-19 (in any position). We identified additional outpatient cases in the EHR via diagnosis codes (see **Supplementary Table 2**) and included telehealth consultations and emergency visits. Exploratory objectives included subgroup analysis for primary and secondary outcomes (COVID-19-related hospitalizations and outpatient visits, respectively) by age group (≥50 years, and ≥65 years).

### Statistical analysis

Using a multivariable logistic model for baseline variables, we calculated propensity score, and estimated weights to minimize the influence of potential confounders that we had identified prior to the analysis (**Supplementary Table 3**). We then used stabilized and truncated weights to re-weight the study sample using a 1% asymmetrical trim, to limit the effects of extreme weights on the study sample and performed weighting separately for the subgroup analysis by age group.

Using Cox regression models, we estimated unadjusted hazard ratios (HR) for the primary and secondary outcomes after propensity score weighting with exposure as the only predictor. Using multivariate Cox regression models, we then estimated the adjusted HRs, with any baseline variable with a standardized mean difference (SMD) of >0.1 after weighting included as a covariate. We considered the cohort to be well balanced if the difference in SMD between the comparison groups was <0.1.

To explore the possibility of residual confounding, we used a negative control. We evaluated the risk of COVID-19-related hospitalization within the first 7 days after vaccination and compared the cumulative incidence among the exposure groups during this immunization lag period. This approach allows us to ensure that both groups are comparable before the cohort entry date (CED).

We calculated rVE as 100× (1−HR) for both unadjusted and adjusted estimates, with 95% confidence intervals (95% CI). To control inflation of type 1 error rate due to multiple testing (family wise error rate), we used a step-down testing procedure for secondary endpoints and the age subgroup analysis by adjusting the rejection criteria for each of the individually tested hypotheses.^41^ We did statistical testing at a significance level of p=0.05 and stopped when a secondary endpoint did not reach statistical significance.

All statistical analyses were performed using SAS 9.4 or R Statistical Software (v4.1.3)^42^ *survival* (v3.2-13).

### Sensitivity analyses

We performed two sensitivity analyses. First, since the main analyses were conducted on closed claims, we compared open versus closed claims. For patients with open claims, follow-up was concluded at the end of data availability. The analysis utilized the last evidence of database activity (EHR or claims) as the endpoint for follow-up (see **Supplementary Materials** for further details).

Additionally, we performed sensitivity analyses using a cohort entry date of 14 days after vaccination (index date) instead of 7 days as specified in the main closed claims analysis.

## Results

We had records available for 8,929,450 individuals who had received a bivalent mRNA vaccine within the study timeframe (**Figure 2**). Of these, 2,748,358 individuals were eligible for inclusion in the pre-weight cohort, with 1,049,575 in the mRNA-1273.222 group and 1,698,783 in the BNT162b2 Bivalent group. After weighting a total of 1,034,538 and 1,670,666 were included in the mRNA-1273.222 and BNT162b2 Bivalent groups respectively (**Table 1**). After weighting, key baseline demographic variables were broadly similar between the two groups: mean age was 58–59 years across groups, 58% were women, and 40% were white (where race was available). The majority (95%) of individuals in both groups had received the bivalent vaccine between September and December 2022 (**Supplementary Figure 1**). Median duration of follow-up was 108 and 107 days for mRNA-1273.222 and BNT162b2 Bivalent vaccines, respectively (**Table 1**). Overall, 69.5% of individuals vaccinated with mRNA-1273.222 and 68.2 % of individuals vaccinated with BNT162b2 Bivalent had underlying medical conditions. As expected, we found a nearly identical hospitalization risk pattern in the evaluation of the negative control for both vaccines, with a p-value of 0.46, indicating that the groups were well balanced. (**Supplementary Figure 2**). Baseline characteristics of the age subgroups are provided in **Supplementary Tables 4 and 5**.

**Figure 2.**
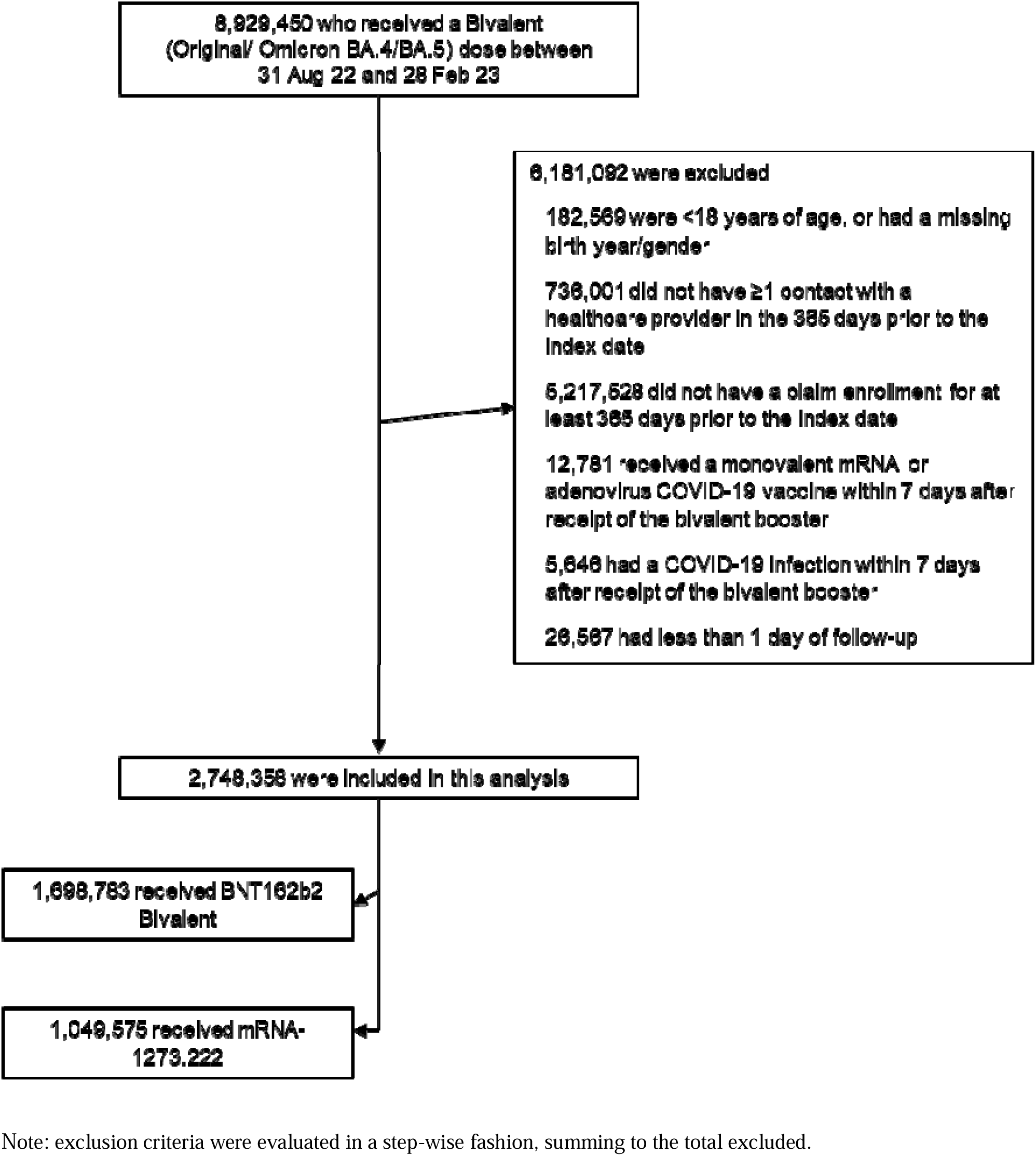
Selection of study populations: bivalent vaccine recipients for inclusion in the analysis.

**Table 1.**
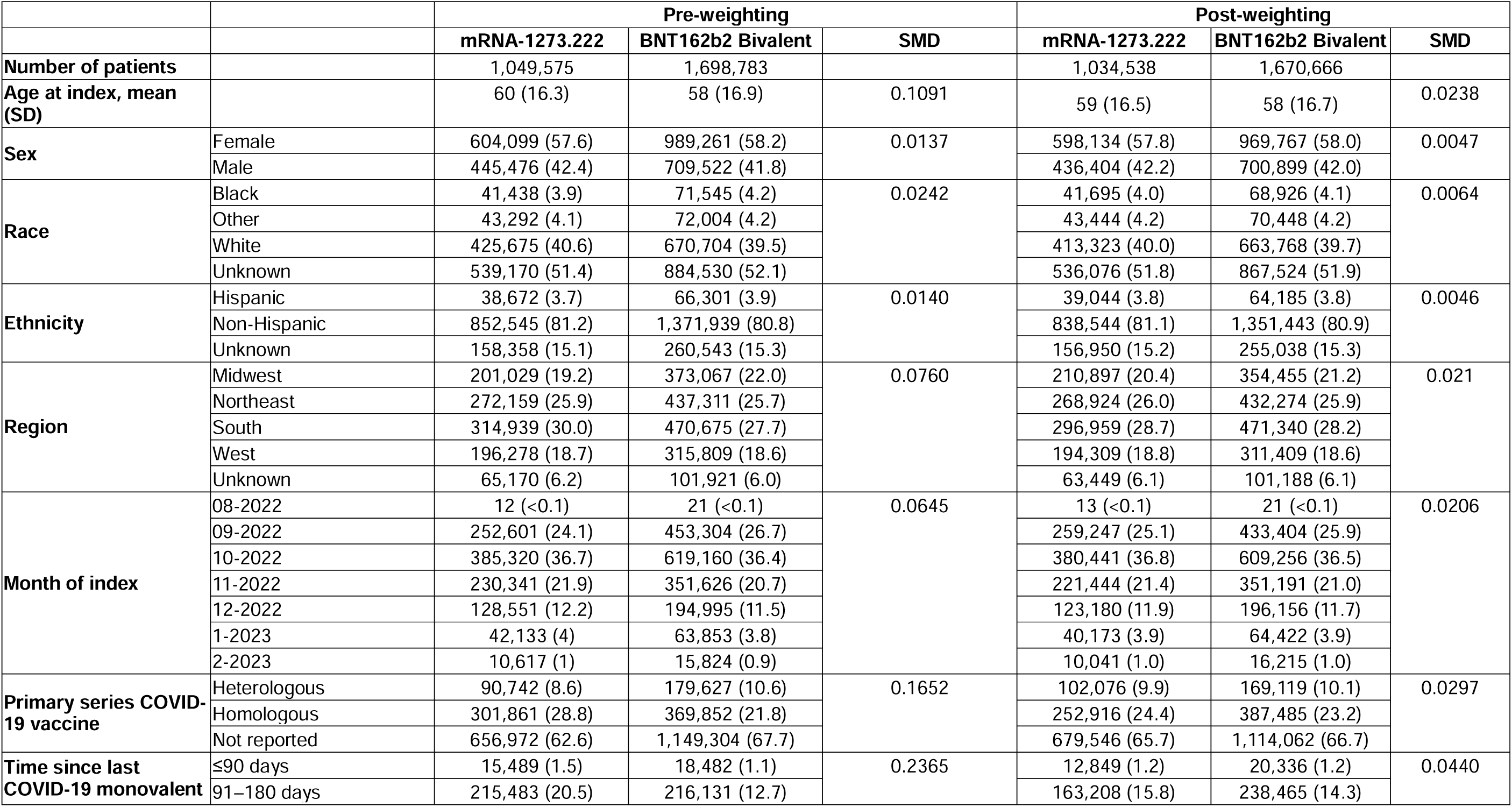
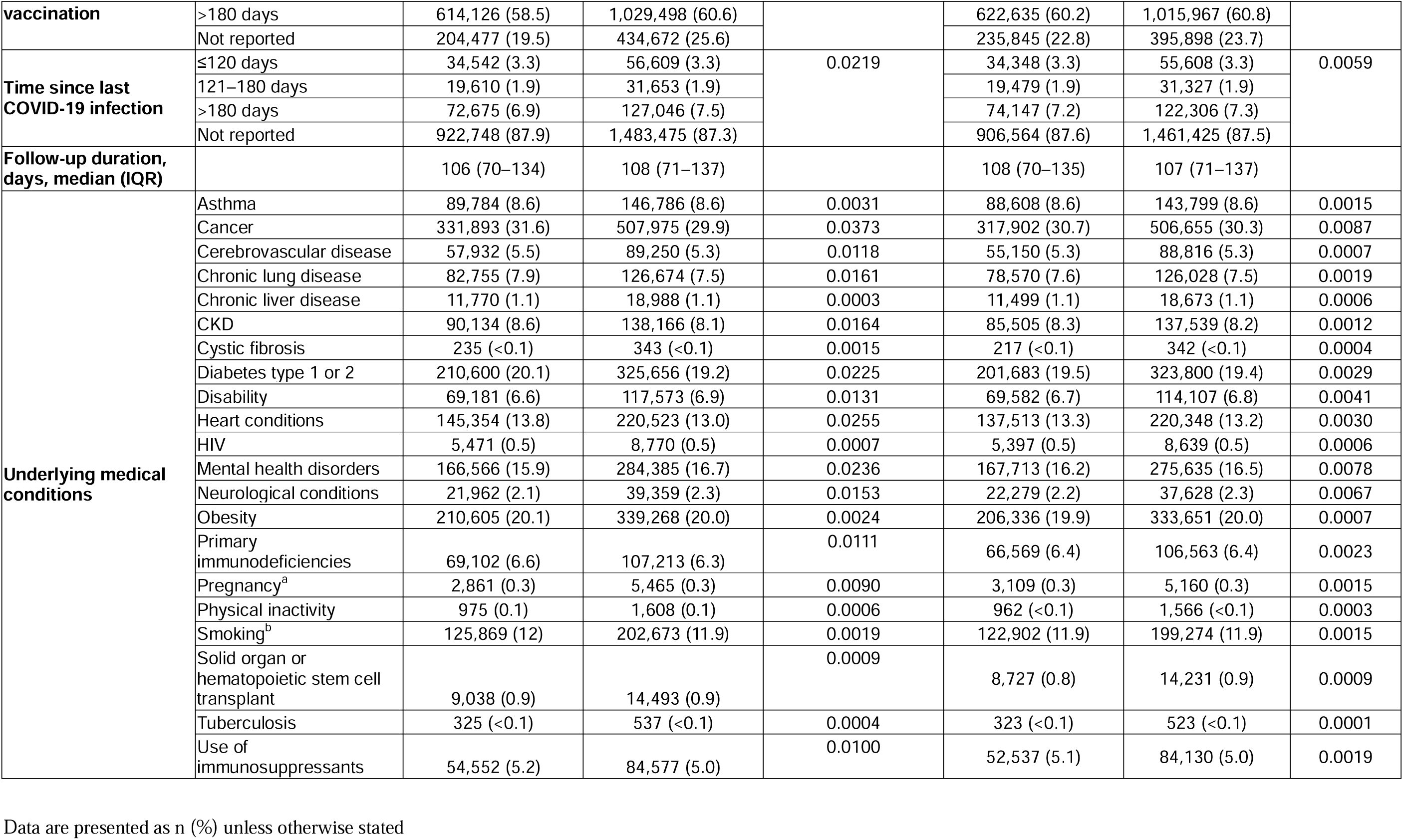
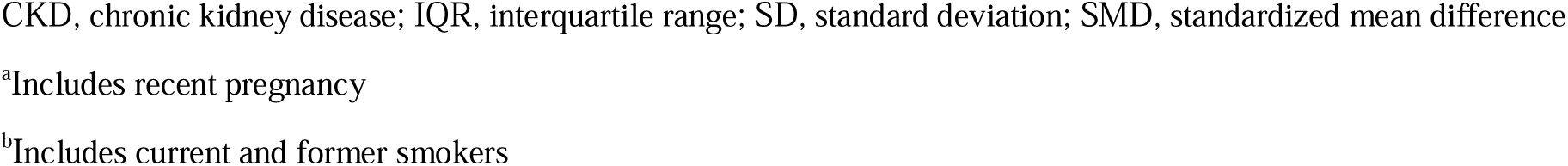
Key baseline characteristics of patients included in the mRNA-1273.222 and BNT162b2 Bivalent vaccine groups (pre- and post-weighting).

### Relative vaccine effectiveness for the overall study population and by age group

Overall, 1,032 (0.10%) individuals who had received mRNA-1273.222 and 1,855 (0.11%) who had received BNT162b2 Bivalent were hospitalized for COVID-19. Of these, 91.2% and 92.5% were ≥50 years of age, and 71.6% and 74.5% were ≥65 years of age at the index date for the two vaccines, respectively. Over three-quarters reported no previous documented COVID-19 infection (78.2% and 76.2% for mRNA-1273.222 and BNT162b2 Bivalent vaccines, respectively), and 49.0% and 47.0% had last received a monovalent COVID-19 vaccine >180 days previously, for the two groups, respectively. The most common underlying medical conditions with increased risk for severe COVID-19 outcomes in hospitalized patients were cancer (mRNA-1273.222: 42.3%; BNT162b2 Bivalent: 44.6%), heart conditions (46.6%; 49.0%), and diabetes (40.4%; 44.0%) (**Supplementary Table 6**).

The rVE against COVID-19-related hospitalizations was 9.8% (95% CI: 2.6%–16.4%; p=0.008) for mRNA-1273.222 compared with BNT162b2 Bivalent vaccines across the entire study population. Differences between vaccines were evident within the first few weeks and persisted over the 6 months after vaccination (**Supplementary Figure 3**). As cohorts were well matched after inverse probability of treatment weighting, no covariates were included in the multivariable models for the overall population; therefore, estimates for both unadjusted and adjusted rVE are equal. Estimates of rVE against COVID-19-related outpatient visits were 5.1% (95% CI: 3.2%– 6.9%; p<0.001) for mRNA-1273.222 compared with the BNT162b2 Bivalent vaccine (**Table 2**).

**Table 2.** Unadjusted and adjusted rVE for the bivalent vaccines (Original/Omicron BA.4/BA.5), mRNA-1273.222 versus BNT162b2 Bivalent for the overall study population.

| COVID-19-related outcome | Unadjusted rVE | Adjusted rVE | P value<sup>a</sup> |
| --- | --- | --- | --- |
| Hospitalization | 9.8% (2.6% – 16.4%) | 9.8% (2.6% – 16.4%) | 0.008 |
| Outpatient | 5.1% (3.2% – 6.9%) | 5.1% (3.2% – 6.9%) | <0.0001 |

rVE, relative vaccine effectiveness
<sup>a</sup>Associated p values are given for the adjusted rVE, with secondary outcomes adjusted for multiple testing.

As the study met both primary and secondary endpoints for the overall population, subgroup analysis was performed by age group. rVE against both COVID-19-related hospitalizations and outpatient visits increased with increasing age, with adjusted rVE against the two outcomes at 11.0% (95% CI: 3.7%–17.7%; p=0.004) and 7.3% (5.3%–9.3%; p<0.0001) for hospitalizations and outpatient visits, respectively, in the aged ≥50 years subgroup, and 13.5% (95% CI: 5.5%–20.8%; p=0.001) and 10.7% (8.2%–13.1%; p<0.0001) in the aged ≥65 years subgroup, respectively (**Table 3**).

**Table 3.** Unadjusted and adjusted rVE for the bivalent vaccines (Original/Omicron BA.4/BA.5), mRNA-1273.222 versus BNT162b2 Bivalent for the subgroups and sensitivity outcomes.

| COVID-19-related Outcome | Unadjusted rVE | Adjusted rVE | P value<sup>a</sup> |
| --- | --- | --- | --- |
| **Subgroup analysis by age** |  |  |  |
| **≥50 years** |  |  |  |
| Hospitalization | 11.0% (3.7% – 17.7%) | 11.0% (3.7% – 17.7%) | 0.004 |
| Outpatient | 7.3% (5.3% – 9.3%) | 7.3% (5.3% – 9.3%) | <0.0001 |
| **≥65 years** |  |  |  |
| Hospitalization | 13.5% (5.5% – 20.8%) | 13.5% (5.5% – 20.8%) | 0.001 |
| Outpatient | 10.7% (8.2% – 13.1%) | 10.7% (8.2% – 13.1%) | <0.0001 |
| **Sensitivity analysis** |  |  |  |
| **Open claims** |  |  |  |
| Hospitalization | 12.9% (9.1% – 16.4%) | 12.9% (9.1% – 16.4%) | <0.0001 |
| Outpatient | 5.8% (4.7% – 6.9%) | 5.8% (4.7% – 6.9%) | <0.0001 |
| **Cohort entry date 14 days post-vaccination closed claims** |  |  |  |
| Hospitalization | 10.6% (3.1% – 17.5%) | 10.6% (3.1% – 17.5%) | 0.006 |
| Outpatient | 4.6% (2.6% – 6.5%) | 4.6% (2.6% – 6.5%) | <0.0001 |

rVE, relative vaccine effectiveness
<sup>a</sup>Associated p values are given for the adjusted rVE, with secondary outcomes adjusted for multiple testing.

### Sensitivity analyses

Point estimates of rVE from evaluation of open claims were similar to those seen in the closed claims analysis (n=3,065,980 for mRNA-1273.222 and n=4,703,189 for BNT162b2 Bivalent): 12.9% (95% CI: 9.1%–16.4%) and 5.8% (4.7%–6.9%), against COVID-19-related hospitalizations and outpatient visits, respectively (**Table 3**).

In addition, extending the time between vaccination (index date) and cohort entry date from 7 to 14 days in the closed claims analysis resulted in rVE estimates of 10.6% (95% CI: 3.1%– 17.5%) and 4.6% (2.6%–6.5%) against COVID-19-related hospitalizations and outpatient visits, respectively (**Table 3**).

## Discussion

This is the first study that directly evaluates the relative vaccine effectiveness of bivalent mRNA vaccines containing the ancestral (original) and omicron BA.4/BA.5 strains in prevention of COVID-19-related hospitalizations and outpatient visits. In this retrospective analysis of real-world data on more than 2.5 million vaccinated individuals, mRNA-1273.222, bivalent vaccine (Original/ Omicron BA.4/BA.5) appears significantly more effective than BNT162b2 bivalent vaccine (Original/ Omicron BA.4/BA.5) in preventing COVID-19-related hospitalizations and outpatient visits in adults. The rVE against both hospitalizations and outpatient visits increased with increasing age. This is particularly important given the disproportionate burden of COVID-19 related morbidity and mortality in older adults.

The results of this study echo those observed following analysis of the primary series and monovalent booster in the same data-set.^32^ Similar to the current study, incremental benefits in rVE against hospitalizations were observed following the monovalent booster with increasing age, with rVE of 40% (95% CI: 11%–60%) in adults ≥65 years and 12% (0%– 30%) in 18 to 64 age group. Other studies have also shown increased effectiveness of mRNA-1273 versus BNT162b2 boosters against symptomatic infections and severe outcomes.^29, 31^ In a matched cohort study comparing the effectiveness of a single monovalent booster dose of mRNA-1273 and BNT162b2 using the OpenSAFELY-TPP research platform in the UK, the hazard ratio of risk of hospital admission at 20 weeks post-booster was 0.89 (95% CI: 0.82 to 0.95), indicating a modest benefit of mRNA-1273.^29^ In a similar matched cohort study of US Veterans, risks of 16-week COVID-19 outcomes were also lower for mRNA-1273 compared with BNT162b2 monovalent booster, with an excess number of COVID-19 hospitalization events over the 16-week post-booster period of 10.6 (95% CI: 5.1–19.7) for BNT162b2 compared with mRNA-1273.^31^ Moreover, while there was no formal comparison between the vaccines, a study evaluating the effectiveness of bivalent vaccines containing any omicron subvariant also showed higher effectiveness of mRNA-1273.222 versus BNT162b2 Bivalent against infection. However, other outcomes were not evaluated in this study.^43^

The observed differences in effectiveness between the mRNA bivalent vaccines, particularly in older adults, may be explained at least in part by differences in vaccine humoral and T cell immunity. In terms of humoral immunity, Al-Sadeq et al., have shown that mRNA-1273 induces a significantly greater antibody immune response compared with the BNT162b2 vaccine.^44^ Others have shown that this difference is consistent against different variants of concern, persisting even 6-8 months following vaccination.^45–48^ Moreover, a greater and more durable immune response with mRNA-1273 compared with BNT162b2 has been demonstrated among older adults. Studies have shown that age is negatively correlated with antibody levels in participants who were vaccinated with BNT162b2 but not among mRNA-1273 vaccinees. Vaccination with mRNA-1273 induced higher antibody levels with a corresponding lower waning rate that was not age-dependent, as opposed to the BNT162b2 cohort in which older individuals generate lower antibody levels with a higher waning ratio when compared to young adults.^46, 49^

Antibodies that can rapidly opsonize pathogens and drive pathogen clearance via opsinophagocytosis or killing are key to protection against many other respiratory pathogens, such as *Streptococcus pneumonia*.^50^ Thus, in addition to neutralizing antibodies, antibodies that bind to viral proteins may also contribute to the immune control of infection through the increased clearance of free virus or by targeting infected cells for immune clearance.^51^ Indeed, there is emerging evidence describing functional antibody differences, beyond neutralization, between the vaccines that may account for differential mucosal protection.

Kaplonek et al evaluated differences in antibody levels and pathogen-killing functions across the two mRNA vaccines. In this study, mRNA-1273 induced more opsonophagocytic and cytotoxic antibodies compared to BNT162b2 that may be important in the rapid capture and clearance of the virus from the mucosal tract.^52^ Furthermore, following, mRNA-1273 vaccination higher levels of IgA were measure systemically compared to BNT162b2 which may be important for mucosal protection aiding the immune response even when new viral variants may emerge that subvert the neutralizing antibody response.^53^ Along these lines, systemic vaccine induced IgA levels have been identified as a strong correlate of protection against viral breakthrough following boosting in the setting of high neutralizing antibody responses.^54^

In addition to the humoral response, several studies have shown that mRNA-1273 vaccination elicited more robust and long-term T-cells responses compared with BNT162b2.^55–58^ Notably, Zhang et al. found that memory CD4+ T cell frequencies were significantly lower following immunization with BNT162b2 compared with mRNA-1273 (between 1.8- and 2.6-fold lower) measured over 6 months.^56^ The enhanced functional humoral immune response and T-cell immunity observed following vaccination with mRNA-1273 may contribute to the differences in effectiveness observed, especially in the older age groups who often have lower responses to immunization in part related to immunosenescence.^59, 60^

In this study, we used real-world data from two integrated sources. Utilization of a comprehensive real-world dataset that integrates various sources of patient information enables the assessment of effectiveness outcomes that are typically not examined in randomized trials. It also facilitates the estimation of effects with robust statistical power. Relying solely on claims data or electronic health records data in many cases cannot offer a complete, accurate, and timely understanding of an individual’s health status. However, combining integrated databases that link both EHR and claims data can provide a well-rounded perspective on an individual’s health status and utilization of health care services.^61^ Moreover, the richness and comprehensiveness of the data in this study allowed the adjustment of well-established confounders using robust confounder adjustment methodology. The information on exposure, outcome, and covariates was retrospectively collected from patient records in a consistent manner across all exposure cohorts, reducing the likelihood of differential misclassification of these elements. By employing EHRs linked to claims data to determine exposure status, the chances of exposure misclassification were minimized as specific product codes were used to identify vaccination status based on the type of vaccine. The conclusions from the main analysis were confirmed by planned sensitivity analyses.

As with all observational or quasi-experimental studies, it also has limitations. First, in this study, we did not evaluate long-term vaccine effectiveness. Our follow-up period for the study was relatively short, with a maximum of 6 months post-booster until the data cut-off. Given the differences in antibody waning and durability observed in immunogenicity studies of earlier vaccine formulations.^17, 45, 62^, it might be that rVE becomes more pronounced over a longer follow-up period. Future analysis, including longer follow-up periods, could provide a broader overview of long-term comparative vaccine performance and help guide decision-making on the optimal interval. In Addition, the current primary analysis was restricted to closed claims, which provide a comprehensive overview of a patient’s healthcare interactions but may not capture all the cases and can be limited in terms of the sample size. However, sensitivity analysis performed on the open claims database supported the findings from the closed claims database. Furthermore, in this study, we included only individuals who had consulted healthcare providers during the previous 365 days, which excluded healthy individuals who received a vaccine but did not use healthcare services during the year prior to receipt, which can also limit our sample size or bias towards more at-risk population. Older adults and those with underlying chronic health conditions, who have an increased risk of more severe COVID-19 outcomes are more likely than average healthy adults to utilize healthcare services^63^. Thus, while our results may not be representative of a population of healthy individuals, they include the most vulnerable population. Finally, the results of the study should be interpreted within the context of its retrospective nature. As with the previous analysis on the primary series and monovalent booster,^32^ there may have been differences between the two vaccine groups that were not fully accounted for by the pre-defined covariates and thereby confounded rVE estimates. It is possible that individuals with greater risk for COVID-19-related severe outcomes were vaccinated with BNT162b2 Bivalent compared to mRNA-1273.222 due to its earlier availability. While this could introduce some residual bias, we do not believe that it is a significant concern. Our negative control test results demonstrated no difference between the groups in term of hospitalizations, suggesting that patient behavior bias may not have influenced our findings. Additionally, the propensity scoring analysis ensured the groups were well-balanced in terms of known comorbidities, reducing the likelihood of differences in patient behavior based on risk factors.

In summary, both bivalent vaccines have been shown to provide substantial protection against both COVID-19-related hospitalizations and outpatient visits. Here we report that the mRNA-1273.222 vaccine (Moderna Bivalent, Original/ Omicron BA.4/BA.5) provided greater protection against hospitalizations and outpatient visits compared with BNT162b2 Bivalent vaccine (Pfizer-BioNTech Bivalent, Original/ Omicron BA.4/BA.5) during a period of omicron BA.4/BA.5 transmission predominance. The results of this study can help guide decision-makers and raise the need to evaluate vaccine differences of current and future formulations to ensure optimized protection against continually emerging SARS-CoV-2 variants and subvariants.

## Acknowledgements

The authors would like to thank the following individuals for their contribution: Editorial assistance was provided by Dr Jenny Engelmoer (Sula Communications BV)

## Data Availability Statement

Individual-level data reported in this study are not publicly shared. Upon request, and subject to review, Veradigm may provide the de-identified aggregate-level data that support the findings of this study. Deidentified data (including participant data as applicable) may be shared upon approval of an analysis proposal and a signed data access agreement. Individual-level data reported in this study shared fully with regulatory agencies.

