## Supplement Material for "Comparative Effectiveness of the Bivalent (Original/Omicron BA.4/BA.5) mRNA COVID-19 Vaccines mRNA-1273.222 and BNT162b2 Bivalent in Adults in the United States"

**Supplementary Materials**

**Supplementary information on the open and closed claims datasets**

A three-tiered definition of closed claims was used to generate a closed claims dataset. First, a claim was closed for a patient if all medical and pharmacy transactions had been received in a given time period. If not all transactions for a patient had been received, the data was considered “open” for that patient. Second, a source was “closed/payer complete” if all patient claims were “closed” under that payer. Third, time periods in which patients are closed could be determined by the enrollment dates; hence enrollment periods were considered closed if the end date of enrollment was closed for that patient. Medical and pharmacy claims included in a closed claims dataset were all post-adjudication claims. Therefore, the closed claims dataset that was used in the main analysis captured all claims associated with a patient (medical and pharmacy), during the time period in which they are enrolled. The open claims analysis used all claims available for a given patient, regardless of whether they were during an enrollment period, sourced from a payer, or considered closed.

**Supplementary Table 1**. List of CVX, CPT, and NDC codes used to identify bivalent (Original/ Omicron BA.4/BA.5) COVID-19 vaccines from the Veradigm EHR and linked claims datasets.

| **Code Type** | **Code** | **Description** | **Vaccine** | **Dose Sequence** |
| --- | --- | --- | --- | --- |
| CPT | 91313 | Severe acute respiratory syndrome coronavirus 2 (SARS-CoV-2) (coronavirus disease [COVID-19]) vaccine, mRNA-LNP, spike protein, bivalent, preservative free, 50 mcg/0.5 mL dosage, for intramuscular use | mRNA-1273.222 | Bivalent booster |
| CPT | 0134A | Immunization administration by intramuscular injection of severe acute respiratory syndrome coronavirus 2 (SARS-CoV-2) (coronavirus disease [COVID-19]) vaccine, mRNA-LNP, spike protein, bivalent, preservative free, 50 mcg/0.5 mL dosage, booster dose | mRNA-1273.222 | Bivalent booster |
| NDC | 80777-0280-05 | mRNA-1273 COVID-19 Vaccine, BL (80777-280-05) | mRNA-1273.222 | Bivalent booster |
| NDC | 80777-0280-99 | mRNA-1273 COVID-19 Vaccine, Bivalent; 0.1 mg/mL; 10 VIAL in 1 CARTON, 2.5 mL in 1 VIAL | mRNA-1273.222 | Bivalent booster |
| CVX | 229 | SARS-COV-2 (COVID-19) vaccine, mRNA, spike protein, LNP, bivalent, preservative free, 50 mcg/0.5 mL dose [Pre-EUA mRNA-1273 bivalent adult booster original strain + omicron strain] | mRNA-1273.222 | Bivalent booster |
| CVX | 300 | SARS-COV-2 (COVID-19) vaccine, mRNA, spike protein, LNP, bivalent booster, preservative free, 30 mcg/0.3 mL dose, tris-sucrose formulation; EUA authorized BNT162b2 adult bivalent booster ages 12+ yrs, original strain + omicron BA.4/BA.5. Not Authorized by WHO. Non-US Tradename for same formulation (Comirnaty Bivalent) counted toward immunity in US | BNT162b2 Bivalent | Bivalent booster |
| CPT | 91312 | Severe acute respiratory syndrome coronavirus 2 (SARS-CoV-2) (coronavirus disease [COVID-19]) vaccine, mRNA-LNP, bivalent spike protein, preservative free, 30 mcg/0.3 mL dosage, tris-sucrose formulation, for intramuscular use | BNT162b2 Bivalent | Bivalent booster |
| CPT | 0124A | Immunization administration by intramuscular injection of severe acute respiratory syndrome coronavirus 2 (SARS-CoV-2) (coronavirus disease [COVID-19]) vaccine, mRNA-LNP, bivalent spike protein, preservative free, 30 mcg/0.3 mL dosage, tris-sucrose formulation, booster dose | BNT162b2 Bivalent | Bivalent booster |
| NDC | 59267-0304-01 | PFIZER COVID BIVALENT BOOST(12Y UP)(ORIG-BA.4/5)(GRAY)(EUA) | BNT162b2 Bivalent | Bivalent booster |
| NDC | 59267-0304-02 | PFIZER COVID BIVALENT BOOST(12Y UP)(ORIG-BA.4/5)(GRAY)(EUA) | BNT162b2 Bivalent | Bivalent booster |
| NDC | 59267-1404-01 | PFIZER COVID BIVALENT BOOST(12Y UP)(ORIG-BA.4/5)(GRAY)(EUA) | BNT162b2 Bivalent | Bivalent booster |
| NDC | 59267-1404-02 | PFIZER COVID BIVALENT BOOST(12Y UP)(ORIG-BA.4/5)(GRAY)(EUA) | BNT162b2 Bivalent | Bivalent booster |

CPT, current procedural terminology; NDC, national drug codes; CVX, vaccine administered code set;

### **Supplementary Table 2**. ICD-10 and SNOMED codes for the outcome of COVID-19-related medical encounters identified from individuals’ primary care EHRs, pharmacy, and medical claims.

| **Code type** | **Codes for COVID-related medical encounters** |
| --- | --- |
| **ICD-10-CM** | J1282, U071, U072, B34.2 |
| **SNOMED** | 1119302008, 119731000146105, 119741000146102, 119751000146104, 119981000146107, 1240411000000107, 1240521000000100, 1240531000000103, 1240541000000107, 1240561000000108, 1240581000000104, 674814021000119106, 840533007, 840534001, 840536004, 840539006, 866151004, 866152006, 870577009, 870588003, 870589006, 870590002, 870591003, 871562009 |

EHR, electronic health records; ICD, international classification of diseases; SNOMED, systematized nomenclature of medicine clinical terms.

**Supplementary Table 3**. List of potential confounding baseline variables which were weighted using propensity weighting scoring prior to data analysis

| **Variable** | **Variable Type** | **Categories** |
| --- | --- | --- |
| **Age^a^** | Categorical | 5-year range from 25 years onwards (i.e., 18–24, 25–29, 30–34, 35–39. etc) |
| **Sex** | Categorical | Female, Male |
| **Race** | Categorical | - Black or African American - White - Other - Not Reported |
| **Ethnicity** | Categorical | - Hispanic - Non-Hispanic - Not Reported |
| **Insurance Type** | Categorical | - Commercial - Medicaid - Medicare Advanced - Medicare FFS - Other - Unknown |
| **Geographic region^b^** | Categorical | - **Northeast** (Connecticut, Maine, Massachusetts, New Hampshire, New Jersey, New York, Pennsylvania, Rhode Island, Vermont ) - **Midwest** (Illinois, Indiana, Iowa, Michigan, Minnesota, Missouri, Nebraska, North Dakota, Ohio, Kansas, South Dakota, Wisconsin) - **South** (Alabama, Arkansas, Delaware, District of Columbia, Florida, Georgia, Kentucky, Louisiana, Maryland, Mississippi, North Carolina, Oklahoma, South Carolina, Tennessee, Texas, Virginia, West Virginia) - **West** (Alaska, Arizona, California, Colorado, Hawaii, Idaho, Montana, Nevada, New Mexico, Oregon, Utah, Washington, Wyoming) - **Other** - **Not Reported/missing** |
| **Underlying medical conditions (with increased risk for severe COVID-19 outcomes^c^ )** | Categorical | - Asthma - Cancer - Cerebrovascular disease - Chronic kidney disease - Chronic lung diseases - Chronic liver disease - Cystic fibrosis - Diabetes mellitus, types 1 and 2 - Disabilities, including Down syndrome - Heart conditions - HIV - Mental health conditions - Neurological conditions - Obesity (BMI >30 kg/m^2^) - Physical inactivity - Pregnancy and recent pregnancy - Primary immunodeficiencies - Smoking, current and former - Solid organ or blood stem cell transplantation - Tuberculosis - Use of corticosteroids or other immunosuppressive medications |
| **Number of outpatient visits in year prior to index date** | Continuous | – |
| **Number of all-cause hospital admission in year prior to index data** | Continuous | – |
| **Data source of recorded vaccination^d^** | Categorical | - Inpatient claim - Outpatient claim - Outpatient EHR data - Pharmacy claim |
| **Primary series vaccination** | Categorical | - Homologous = same brand as bivalent dose - Heterologous = different brand from bivalent dose - Not documented |
| **Time since last COVID-19 monovalent vaccination** | Categorical | - Less than 3 months - 3 to 6 months - 6 months or more - Not documented |
| **Time since last COVID-19 infection** | Categorical | - Less than 4 months - 4 to 6 months - 6 months or more - Not documented |
| **Month of index date** | Categorical | Calendar month of index date |

BMI, body mass index; CDC, centers for disease control and prevention; EHR, electronic health records; FFS, fee for service.

^a^Participant age was capped at ≤89 years to prevent possible patient re-identification

^b^Categorized into one of five mutually exclusive geographic locations in the U.S. based on regional census divisions (<https://www2.census.gov/geo/pdfs/maps-data/maps/reference/us_regdiv.pdf>).

^c^As per CDC guidelines: <https://www.cdc.gov/coronavirus/2019-ncov/hcp/clinical-care/underlyingconditions.html#print>.

^d^Counts of patients by recorded vaccination source for each vaccine type are reported. These counts were mutually exclusive and decided in a hierarchical manner.

**Supplementary Table 4**. Baseline characteristics (post-weighting) of individuals ≥50 years vaccinated with mRNA-1273.222 or BNT162b2 Bivalent vaccine.

|  | |  | **mRNA-1273.222** | **BNT162b2 Bivalent** | **SMD** |
| --- | --- | --- | --- | --- | --- |
| **Number of patients** | |  | 773,586 | 1,178,604 |  |
| **Age at index, mean (SD)** | |  | 67 (9.8) | 67 (9.8) | 0.0096 |
| **Sex** | | Female | 441,890 (57.1) | 674,578 (57.2) | 0.0023 |
|  |  | Male | 331,696 (42.9) | 504,026 (42.8) |  |
| **Race** | | Black | 34,192 (4.4) | 53,214 (4.5) | 0.0048 |
|  |  | Other | 28,091 (3.6) | 42,554 (3.6) |  |
|  |  | White | 330,585 (42.7) | 502,282 (42.6) |  |
|  |  | Unknown | 380,720 (49.2) | 580,554 (49.3) |  |
| **Ethnicity** | | Hispanic | 27,889 (3.6) | 43,003 (3.6) | 0.0037 |
|  |  | Non-Hispanic | 628,014 (81.2) | 955,114 (81.0) |  |
|  |  | Unknown | 117,684 (15.2) | 180,488 (15.3) |  |
| **Geographic region** | | Midwest | 154,244 (19.9) | 246,754 (20.9) | 0.0257 |
|  |  | Northeast | 210,073 (27.2) | 318,540 (27.0) |  |
|  |  | South | 228,525 (29.5) | 340,656 (28.9) |  |
|  |  | West | 133,783 (17.3) | 202,010 (17.1) |  |
|  |  | Unknown | 46,961 (6.1) | 70,645 (6.0) |  |
| **Month of index date** | | 08-2022 | 7 (<0.1) | 10 (<0.1) | 0.0281 |
|  |  | 09-2022 | 193,716 (25.0) | 309,209 (26.2) |  |
|  |  | 10-2022 | 293,896 (38.0) | 443,231 (37.6) |  |
|  |  | 11-2022 | 159,921 (20.7) | 237,589 (20.2) |  |
|  |  | 12-2022 | 88,664 (11.5) | 132,321 (11.2) |  |
|  |  | 1-2023 | 29,837 (3.9) | 44,812 (3.8) |  |
|  |  | 2-2023 | 7,545 (1.0) | 11,433 (1.0) |  |
| **Primary series COVID-19 vaccine** | | Heterologous | 65,270 (8.4) | 108,088 (9.2) | 0.0410 |
|  |  | Homologous | 184,827 (23.9) | 263,818 (22.4) |  |
|  |  | Not reported | 523,490 (67.7) | 806,699 (68.4) |  |
| **Time since last COVID-19 monovalent vaccination** | | ≤90 days | 11,598 (1.5) | 17,290 (1.5) | 0.0400 |
|  |  | 91–180 days | 159,810 (20.7) | 224,925 (19.1) |  |
|  |  | >180 days | 418,407 (54.1) | 649,236 (55.1) |  |
|  |  | Not reported | 183,772 (23.8) | 287,154 (24.4) |  |
| **Time since last COVID-19 infection** | | ≤120 days | 26,820 (3.5) | 41,276 (3.5) | 0.0084 |
|  |  | 121–180 days | 14,095 (1.8) | 21,416 (1.8) |  |
|  |  | >180 days | 55,039 (7.1) | 86,306 (7.3) |  |
|  |  | Not reported | 677,633 (87.6) | 10,29,606 (87.4) |  |
| **Underlying medical conditions** | | Asthma | 66,720 (8.6) | 102,181 (8.7) | 0.0016 |
|  |  | Cancer | 276,033 (35.7) | 418,297 (35.5) | 0.0040 |
|  |  | Cerebrovascular disease | 54,803 (7.1) | 84,117 (7.1) | 0.0021 |
|  |  | Chronic lung disease | 77,622 (10.0) | 118,678 (10.1) | 0.0012 |
|  |  | Chronic liver disease | 10,348 (1.3) | 15,948 (1.4) | 0.0013 |
|  |  | CKD | 85,312 (11) | 130,593 (11.1) | 0.0017 |
|  |  | Cystic fibrosis | 100 (<0.1) | 141 (<0.1) | 0.0008 |
|  |  | Diabetes type 1 or 2 | 189,989 (24.6) | 290,136 (24.6) | 0.0013 |
|  |  | Disability | 38,177 (4.9) | 58,560 (5.0) | 0.0015 |
|  |  | Heart conditions | 137,395 (17.8) | 209,653 (17.8) | 0.0007 |
|  |  | HIV | 4,145 (0.5) | 6,349 (0.5) | 0.0004 |
|  |  | Mental health disorders | 116,509 (15.1) | 181,524 (15.4) | 0.0095 |
|  |  | Neurological conditions | 22,418 (2.9) | 36,092 (3.1) | 0.0097 |
|  |  | Obesity | 167,583 (21.7) | 257,071 (21.8) | 0.0036 |
|  |  | Primary immunodeficiencies | 57,071 (7.4) | 86,869 (7.4) | 0.0003 |
|  |  | Pregnancy^a^ | 15 (<0.1) | 23 (<0.1) | 0.0001 |
|  |  | Physical inactivity | 786 (0.1) | 1,215 (0.1) | 0.0005 |
|  |  | Smoking^b^ | 109,796 (14.2) | 168,895 (14.3) | 0.0039 |
|  |  | Solid organ or hematopoietic stem cell transplant | 8,011 (1.0) | 12,374 (1.0) | 0.0014 |
|  |  | Tuberculosis | 259 (<0.1) | 395 (<0.1) | 0.000 |
|  |  | Use of immunosuppressants | 45,087 (5.8) | 68,657 (5.8) | 0.0001 |

Data are presented as n (%) unless otherwise stated

CKD, chronic kidney disease; SD, standard deviation; SMD, standardized mean difference.

^a^Includes recent pregnancy

^b^Includes current and former smokers

**Supplementary Table 5**. Baseline characteristics (post-weighting) of individuals ≥65 years vaccinated with mRNA-1273.222 or BNT162b2 Bivalent vaccine.

|  | |  | **mRNA-1273.222** | **BNT162b2 Bivalent** | **SMD** |
| --- | --- | --- | --- | --- | --- |
| **Number of patients** | |  | 445,994 | 641,195 |  |
| **Age at index, mean (SD)** | |  | 74 (6.6) | 74 (6.7) | 0.0023 |
| **Sex** | | Female | 253,468 (56.8) | 365,297 (57.0) | 0.0028 |
|  |  | Male | 192,525 (43.2) | 275,898 (43.0) |  |
| **Race** | | Black | 19,132 (4.3) | 27,806 (4.3) | 0.0030 |
|  |  | Other | 13,813 (3.1) | 19,675 (3.1) |  |
|  |  | White | 204,836 (45.9) | 294,383 (45.9) |  |
|  |  | Unknown | 208,213 (46.7) | 299,331 (46.7) |  |
| **Ethnicity** | | Hispanic | 14,184 (3.2) | 20,630 (3.2) | 0.0031 |
|  |  | Non-Hispanic | 365,189 (81.9) | 524,332 (81.8) |  |
|  |  | Unknown | 66,621 (14.9) | 96,233 (15.0) |  |
| **Geographic region** | | Midwest | 88,084 (19.8) | 132,347 (20.6) | 0.0231 |
|  |  | Northeast | 133,004 (29.8) | 190,573 (29.7) |  |
|  |  | South | 130,925 (29.4) | 184,547 (28.8) |  |
|  |  | West | 67,466 (15.1) | 95,983 (15.0) |  |
|  |  | Unknown | 26,514 (5.9) | 37,745 (5.9) |  |
| **Month of index date** | | 08-2022 | 4 (<0.1) | 5 (<0.1) | 0.0217 |
|  |  | 09-2022 | 121,214 (27.2) | 180,378 (28.1) |  |
|  |  | 10-2022 | 170,795 (38.3) | 243,450 (38) |  |
|  |  | 11-2022 | 85,888 (19.3) | 120,926 (18.9) |  |
|  |  | 12-2022 | 46,044 (10.3) | 65,199 (10.2) |  |
|  |  | 1-2023 | 17650 (4.0) | 25,039 (3.9) |  |
|  |  | 2-2023 | 4,399 (1.0) | 6,198 (1.0) |  |
| **Primary series COVID-19 vaccine** | | Heterologous | 23,370 (5.2) | 43,556 (6.8) | 0.0764 |
|  |  | Homologous | 88,844 (19.9) | 115,802 (18.1) |  |
|  |  | Not reported | 333,781 (74.8) | 481,837 (75.1) |  |
| **Time since last COVID-19 monovalent vaccination** | | ≤90 days | 6,827 (1.5) | 9,565 (1.5) | 0.0418 |
|  |  | 91–180 days | 102,382 (23.0) | 136,674 (21.3) |  |
|  |  | >180 days | 203,504 (45.6) | 302,506 (47.2) |  |
|  |  | Not reported | 133,281 (29.9) | 192,450 (30.0) |  |
| **Time since last COVID-19 infection** | | ≤120 days | 15,150 (3.4) | 22,238 (3.5) | 0.0103 |
|  |  | 121–180 days | 7,257 (1.6) | 10,493 (1.6) |  |
|  |  | >180 days | 28,855 (6.5) | 42,960 (6.7) |  |
|  |  | Not reported | 394,732 (88.5) | 565,504 (88.2) |  |
| **Underlying medical conditions** | | Asthma | 36,304 (8.1) | 52,433 (8.2) | 0.0014 |
|  |  | Cancer | 173,386 (38.9) | 248,505 (38.8) | 0.0025 |
|  |  | Cerebrovascular disease | 44,071 (9.9) | 64,071 (10.0) | 0.0037 |
|  |  | Chronic lung disease | 59,176 (13.3) | 86,016 (13.4) | 0.0043 |
|  |  | Chronic liver disease | 5,852 (1.3) | 8,542 (1.3) | 0.0018 |
|  |  | CKD | 71,141 (16) | 103,363 (16.1) | 0.0046 |
|  |  | Cystic fibrosis | 48 (<0.1) | 67 (<0.1) | 0.0003 |
|  |  | Diabetes type 1 or 2 | 125,880 (28.2) | 181,558 (28.3) | 0.0020 |
|  |  | Disability | 19,111 (4.3) | 27,624 (4.3) | 0.0011 |
|  |  | Heart conditions | 107,398 (24.1) | 155,468 (24.2) | 0.0039 |
|  |  | HIV | 1,217 (0.3) | 1,766 (0.3) | 0.0005 |
|  |  | Mental health disorders | 64,813 (14.5) | 95,743 (14.9) | 0.0113 |
|  |  | Neurological conditions | 21,113 (4.7) | 32,188 (5) | 0.0133 |
|  |  | Obesity | 92,563 (20.8) | 133,914 (20.9) | 0.0032 |
|  |  | Primary immunodeficiencies | 33,885 (7.6) | 48,689 (7.6) | 0.0002 |
|  |  | Physical inactivity | 498 (0.1) | 734 (0.1) | 0.0009 |
|  |  | Smoking^a^ | 68,883 (15.4) | 100,317 (15.6) | 0.0055 |
|  |  | Solid organ or hematopoietic stem cell transplant | 5,561 (1.2) | 8,054 (1.3) | 0.0008 |
|  |  | Tuberculosis | 167 (<0.1) | 237 (<0.1) | 0.0003 |
|  |  | Use of immunosuppressants | 27,115 (6.1) | 38,984 (6.1) | 0.0000 |

Data are presented as n (%) unless otherwise stated.

CKD, chronic kidney disease; SD, standard deviation; SMD, standardized mean difference.

^a^Includes current and former smokers.

**Supplementary Table 6**. Baseline characteristics (post-weighting) of individuals hospitalized for COVID-19, vaccinated with mRNA-1273.222 or BNT162b2 Bivalent vaccine.

|  | |  | **mRNA-1273.222** | **BNT162b2 Bivalent** | **SMD** |
| --- | --- | --- | --- | --- | --- |
| **Number of patients** | |  | 1,032 | 1,855 |  |
| **Age at index, mean (SD)** | |  | 71 (14.0) | 72 (14.2) | 0.0837 |
| **Sex** | | Female | 524 (50.8) | 960 (51.8) | 0.0192 |
|  |  | Male | 508 (49.2) | 895 (48.2) |  |
| **Race** | | Black | 51 (5.0) | 84 (4.5) | 0.1134 |
|  |  | Other | 33 (3.2) | 42 (2.3) |  |
|  |  | White | 426 (41.3) | 693 (37.4) |  |
|  |  | Unknown | 522 (50.5) | 1036 (55.9) |  |
| **Ethnicity** | | Hispanic | 42 (4.1) | 60 (3.2) | 0.0452 |
|  |  | Non-Hispanic | 872 (84.5) | 1586 (85.5) |  |
|  |  | Unknown | 119 (11.5) | 210 (11.3) |  |
| **Geographic region** | | Midwest | 207 (20.1) | 418 (22.5) | 0.089 |
|  |  | Northeast | 327 (31.7) | 589 (31.8) |  |
|  |  | South | 276 (26.8) | 435 (23.5) |  |
|  |  | West | 139 (13.5) | 251 (13.5) |  |
|  |  | Unknown | 83 (8.0) | 162 (8.7) |  |
| **Month of index date** | | 08-2022 | 0 (0.0) | 0 (0.0) | 0.0965 |
|  |  | 09-2022 | 334 (32.4) | 665 (35.9) |  |
|  |  | 10-2022 | 452 (43.8) | 788 (42.5) |  |
|  |  | 11-2022 | 190 (18.4) | 290 (15.7) |  |
|  |  | 12-2022 | 47 (4.6) | 97 (5.2) |  |
|  |  | 1-2023 | 9 (0.8) | 14 (0.8) |  |
|  |  | 2-2023 | 0 (0.0) | 0 (0.0) |  |
| **Primary series COVID-19 vaccine** | | Heterologous | 75 (7.3) | 172 (9.3) | 0.0719 |
|  |  | Homologous | 201 (19.4) | 354 (19.1) |  |
|  |  | Not reported | 756 (73.3) | 1329 (71.7) |  |
| **Time since last COVID-19 monovalent vaccination** | | ≤90 days | 19 (1.8) | 45 (2.4) | 0.0571 |
|  |  | 91–180 days | 224 (21.7) | 401 (21.6) |  |
|  |  | >180 days | 506 (49.0) | 872 (47.0) |  |
|  |  | Not reported | 283 (27.4) | 537 (29.0) |  |
| **Time since last COVID-19 infection** | | ≤120 days | 105 (10.2) | 219 (11.8) | 0.0643 |
|  |  | 121–180 days | 20 (1.9) | 29 (1.6) |  |
|  |  | >180 days | 100 (9.7) | 193 (10.4) |  |
|  |  | Not reported | 807 (78.2) | 1414 (76.2) |  |
| **Underlying medical conditions** | | Asthma | 173 (16.8) | 277 (14.9) | 0.0497 |
|  |  | Cancer | 437 (42.3) | 828 (44.6) | 0.0457 |
|  |  | Cerebrovascular disease | 204 (19.8) | 390 (21.0) | 0.0309 |
|  |  | Chronic lung disease | 326 (31.5) | 655 (35.3) | 0.0796 |
|  |  | Chronic liver disease | 39 (3.8) | 74 (4.0) | 0.0094 |
|  |  | CKD | 343 (33.2) | 640 (34.5) | 0.0265 |
|  |  | Diabetes type 1 or 2 | 417 (40.4) | 817 (44.0) | 0.0748 |
|  |  | Disability | 106 (10.3) | 182 (9.8) | 0.0153 |
|  |  | Heart conditions | 481 (46.6) | 909 (49.0) | 0.0477 |
|  |  | HIV | 9 (0.9) | 12 (0.6) | 0.0242 |
|  |  | Mental health disorders | 287 (27.8) | 554 (29.8) | 0.0445 |
|  |  | Neurological conditions | 137 (13.2) | 274 (14.8) | 0.0448 |
|  |  | Obesity | 304 (29.5) | 571 (30.8) | 0.0278 |
|  |  | Primary immunodeficiencies | 155 (15.1) | 279 (15.0) | 0.0002 |
|  |  | Pregnancy^a^ | 30 (2.9) | 38 (2.1) | 0.0559 |
|  |  | Physical inactivity | 4 (0.4) | 2 (<0.1) | 0.0611 |
|  |  | Smoking^b^ | 303 (29.4) | 568 (30.6) | 0.0263 |
|  |  | Solid organ or hematopoietic stem cell transplant | 67 (6.5) | 139 (7.5) | 0.0380 |
|  |  | Tuberculosis | 1 (0.1) | 4 (0.2) | 0.0147 |
|  |  | Use of immunosuppressants | 118 (11.4) | 200 (10.8) | 0.0191 |

Data are presented as n (%) unless otherwise stated.

^a^Includes recent pregnancy. ^b^Includes current and former smokers.

CKD, chronic kidney disease; SD, standard deviation; SMD, standardized mean difference.

**Supplementary Figure 1.** Percentage of individuals included in the weighted analysis by month of vaccination.

Percentage of recepients

| Month | mRNA-1273.222 | BNT162b2 Bivalent |
| --- | --- | --- |
| 2022-08-01 | 0.0012566 | 0.00125698 |
| 2022-09-01 | 25.0592 | 25.942 |
| 2022-10-01 | 36.774 | 36.4679 |
| 2022-11-01 | 21.4051 | 21.021 |
| 2022-12-01 | 11.9068 | 11.7412 |
| 2023-01-01 | 3.88318 | 3.85607 |
| 2023-02-01 | 0.970578 | 0.970571 |

**Supplementary Figure 2.** Negative control Kaplan-Meier curves for COVID-19–related hospitalization rates 7 days prior to CED in recipients of mRNA-1273.222 or BNT162b2 bivalent vaccine

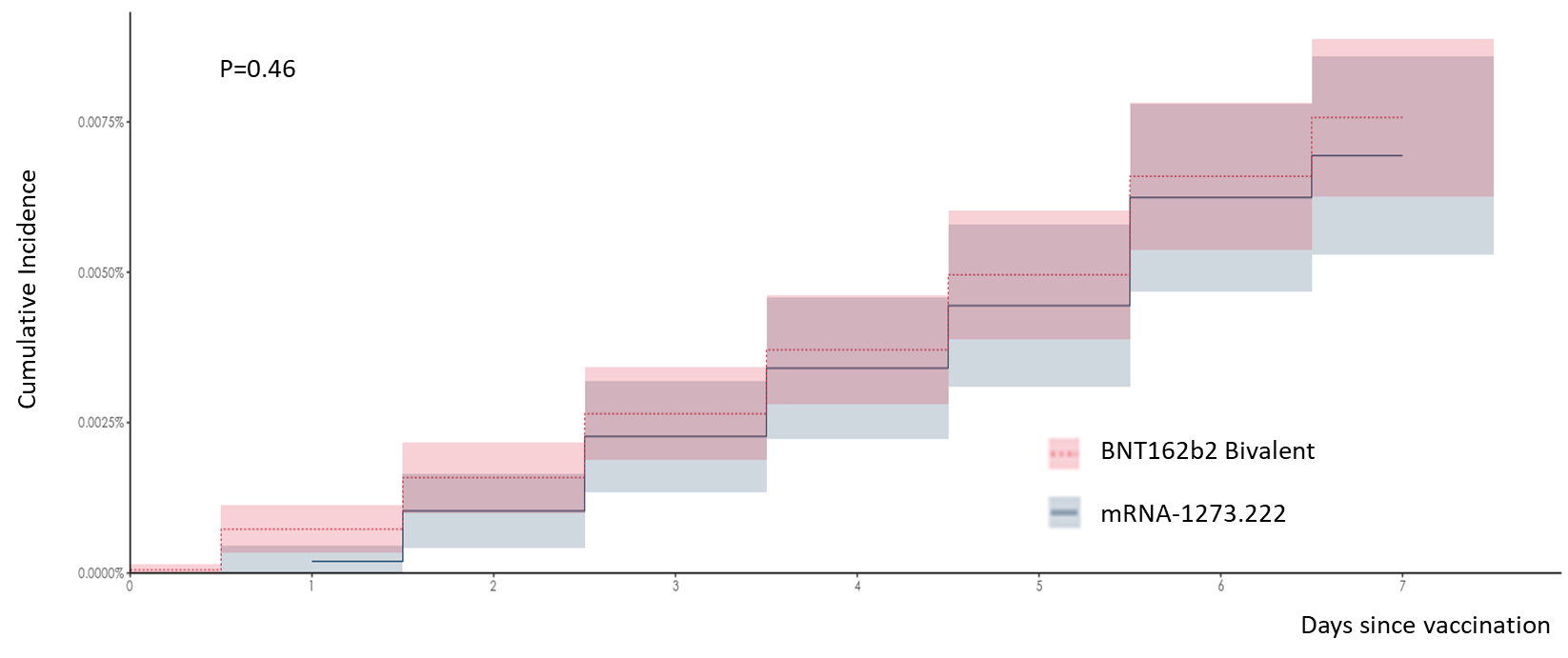

The dashed pink line represents the risk curve for BNT162b2 Bivalent. The solid blue line represents the risk curve for mRNA-1273-222. Shaded areas represent 95% confidence intervals

**Supplementary Figure 3.** Kaplan-Meier curves of COVID-19–related hospitalizations over time in recipients of mRNA-1273.222 or BNT162b2 Bivalent vaccine

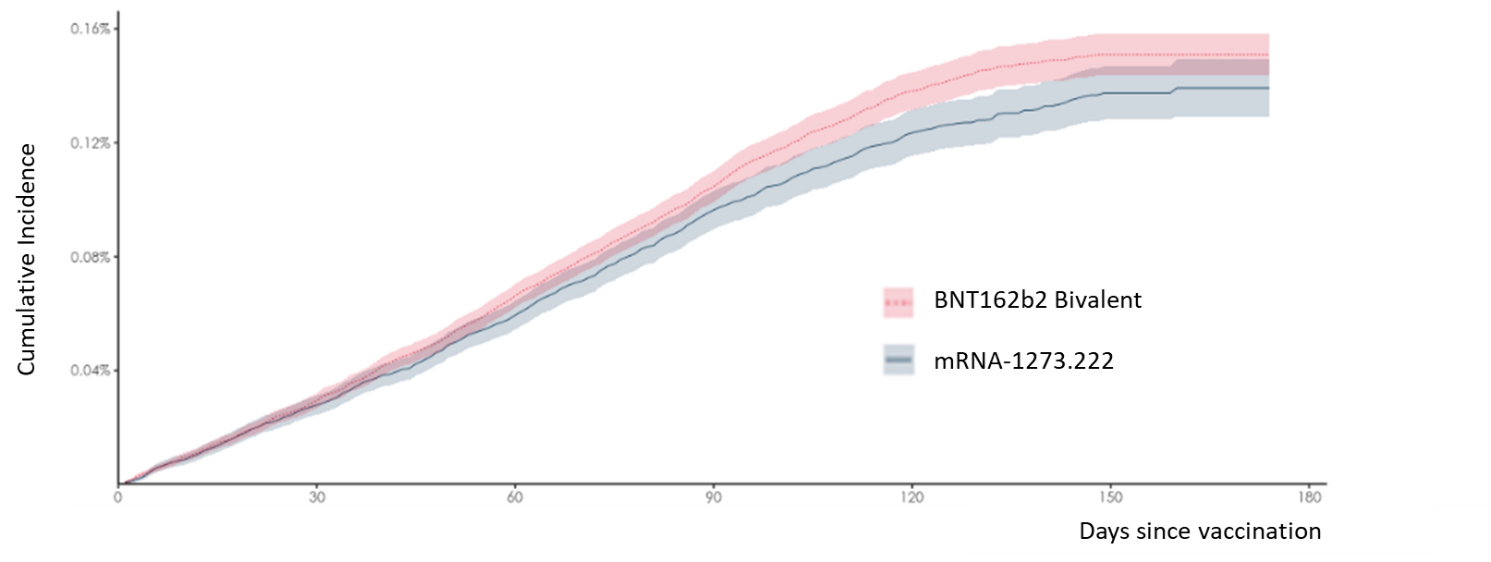

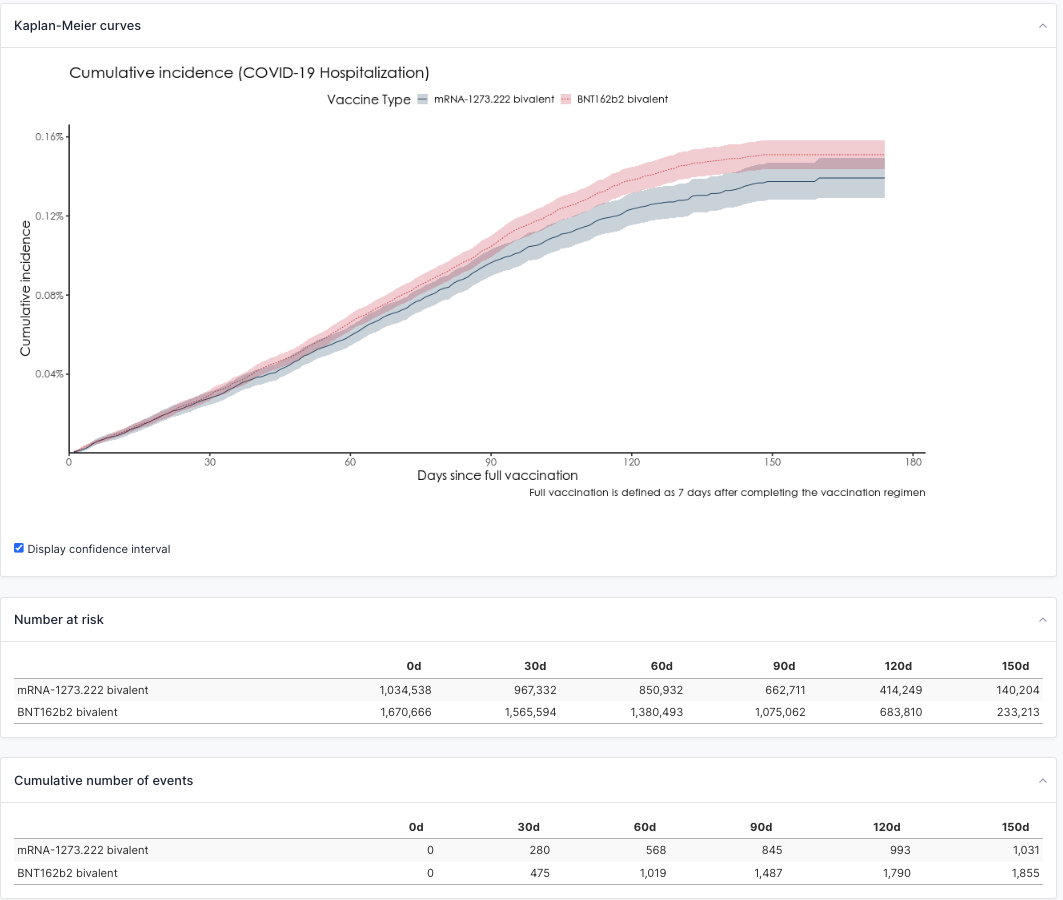
